# Hand hygiene knowledge and practices among health care workers in Uganda: Evidence from Mukono and Kagadi districts

**DOI:** 10.64898/2026.10.06.26364922

**Authors:** Fred Twinomugisha, Richard K. Mugambe, John Bosco Isunju, Esther Buregyeya, David Musoke, Sadiq Murungi, Henry Kajumbula, Habib Yakubu, Richard Kimwanga, Sheillah Simiyu, Raj Suraja, Edine Mliva, Edgar Chilanzi Mulwanda, Musa Manga, Christine L. Moe

**Affiliations:** Makerere University School of Public Health, Department of Disease Control and Environmental Health, Kampala, Uganda; Infectious Diseases Institute, College of Health Sciences, Makerere University, Kampala; Makerere University School of Biomedical Sciences, Department of Microbiology, Kampala, Uganda; The Center for Global Safe Water, Sanitation, and Hygiene, Emory University, Rollins School of Public Health, Atlanta, GA, USA; College of Engineering, University of Dar-es-Salaam, P.O. Box 35131, Dar-es-Salaam, Tanzania; Population Dynamics and Urbanization, African Population and Health Research Center, Nairobi, Kenya; Center of Global Safe Water, Sanitation, and Hygiene, Hubert Department of Global Health, Rollins School of Public Health, Emory University, Atlanta, Georgia; Pan African Consortium of Experts; Zambia Institute of Environmental Health, Lusaka, Zambia; The Water Institute at UNC, Department of Environmental Sciences and Engineering, Gillings School of Global Public Health, University of North Carolina at Chapel Hill, Chapel Hill, NC, USA

**Keywords:** Hand hygiene, Healthcare workers, Knowledge, Alcohol-based hand rub, Uganda, Practice, Infection prevention and control

## Abstract

**Background:** Hand hygiene (HH) is one of the most effective measures to prevent healthcare-associated infections (HAIs) and limit the transmission of antimicrobial-resistant pathogens in healthcare facilities (HCFs). Effective HH programs depend not only on the availability of essential resources but also on healthcare workers’ (HCWs) knowledge, attitudes, and adherence to recommended HH practices. Despite the critical role of these factors in improving infection prevention and control (IPC), evidence on HH knowledge and practices among HCWs remains limited. In Uganda, such evidence from lower-level district health facilities is particularly scarce. This study assessed HH knowledge and practices, along with associated factors, among HCWs at selected health facilities in the Mukono and Kagadi districts of Uganda.

**Methods:** A cross-sectional study was conducted among 586 HCWs at selected health facilities in the Mukono and Kagadi districts from May to June 2025. Data were collected using a structured questionnaire administered via KoboCollect, covering participants’ socio-demographic and professional characteristics, HH knowledge, self-reported HH practices, and facility-related factors. Descriptive statistics summarized the data, and modified Poisson regression with generalized estimating equations (GEE), accounting for clustering of HCWs within facilities, was used to estimate prevalence ratios for factors associated with HH knowledge and practices.

**Results:** Overall, 82.1% of HCWs had adequate HH knowledge, and 43.0% had good HH practice. Nearly all respondents correctly identified the recommended handwashing duration (98.8%), the minimum effective alcohol concentration for alcohol-based hand rub (ABHR) (98.8%), proper HH as the most effective method for preventing HAIs (97.6%), and 89.9% correctly listed all five WHO Moments for HH. HCWs in Mukono were more likely to have adequate HH knowledge than those in Kagadi (aPR = 1.36; 95% CI: 1.15–1.60). HCWs with 6-10 years of experience had higher knowledge of HH than those with less than two years of experience (aPR=1.16, 95% CI: 1.02–1.33). Good HH practice was significantly more common among clinical officers (aPR=1.36, 95% CI: 1.06–1.75) and other cadres (aPR=1.65, 95% CI: 1.14–2.39) than nurses and nursing assistants.

**Conclusion:** HCWs demonstrated high HH knowledge but suboptimal HH practice, suggesting a gap between knowledge and self-reported practice. Interventions should move beyond knowledge dissemination to strengthen compliance through multimodal, cadre-specific strategies, particularly for nurses and nursing assistants.

## Introduction

Hand hygiene (HH) is universally recognized as the single most effective, simple, and cost-effective intervention for preventing healthcare-associated infections (HAIs) and interrupting the transmission of antimicrobial-resistant (AMR) pathogens in healthcare facilities (HCFs) (1–4). HAIs remain a major global public health concern, affecting hundreds of millions of patients annually and contributing substantially to preventable morbidity, mortality, prolonged hospital stays, disability, and escalating healthcare costs (5–7). This burden is compounded by the rise of antimicrobial resistance (8, 9). The burden is disproportionately higher in low-and middle-income countries (LMICs), where the risk of acquiring HAIs is estimated to be two to three times greater than in high-income countries because of constrained healthcare resources, inadequate Infection Prevention and Control (IPC) systems, overcrowded facilities, and limited access to water, sanitation, and hygiene (WASH) services (10, 11). In addition to increasing the burden of HAIs, poor HH practices facilitate the spread of AMR pathogens, undermining the effectiveness of antimicrobial therapy and threatening progress toward universal health coverage and patient safety (12, 13). Consequently, improving HH has become a global priority and a cornerstone of IPC strategies to reduce HAIs, combat AMR, and improve the quality of healthcare delivery.

The World Health Organization (WHO) recommends adherence to the “Five Moments for HH” and the appropriate use of alcohol-based hand rubs (ABHRs) or handwashing with soap and water as fundamental components of comprehensive IPC programs (14–17). Poor compliance has been attributed to multiple interacting factors, including inadequate knowledge, limited availability of hand hygiene infrastructure and supplies, heavy workloads, high patient-to-provider ratios, insufficient managerial support, inadequate supervision, and organizational cultures that do not consistently prioritize patient safety (18–20). Further, although adequate HH knowledge is considered an essential prerequisite for improving compliance, growing evidence suggests that knowledge alone is insufficient to sustain appropriate HH practices (1). Sustained compliance is influenced by multiple interacting factors, including HCWs’ attitudes and perceptions; the availability and accessibility of HH resources; workload and environmental conditions; social and institutional norms; feedback and reminders; and organizational commitment to IPC (21). These factors underscore the importance of addressing both individual and system-level determinants when designing interventions to improve and sustain HH practices.

In Uganda, strengthening HH has become a national priority as part of broader efforts to improve patient safety, reduce HAIs, combat AMR, and enhance the quality of healthcare services (22, 23). The Ministry of Health has adopted the WHO IPC guidelines and implemented several initiatives, including establishing IPC committees, building healthcare workers’ capacity, implementing the WHO multimodal HH improvement strategy, and integrating IPC into national quality improvement programs (24). These efforts have gained further importance following recent outbreaks of Ebola virus disease, COVID-19, Marburg virus disease, cholera, and Mpox, which underscored the critical role of effective HH in protecting both patients and healthcare workers from healthcare-associated transmission (25).

Previous studies in Uganda have examined HH knowledge among HCWs across different healthcare settings and have generally shown varying levels of understanding of HH’s importance and the recommended moments for HH (1). Some studies have reported relatively high levels of knowledge among HCWs, particularly regarding HH’s role in preventing HAIs, while others have identified important gaps in knowledge of the WHO-recommended HH moments and appropriate HH procedures (1, 22). Studies have also shown that HH knowledge may vary across HCW cadres and facility levels, suggesting that context influences knowledge (1). Although several studies in Uganda have assessed HH knowledge, compliance, or broader IPC practices, the available evidence remains fragmented, geographically limited, and largely confined to tertiary hospitals, specific HCW cadres, or individual HCFs (1).

Despite the critical role of HH in HAIs and curbing the spread of AMR, evidence on HH knowledge and practices among HCWs in Uganda remains limited and fragmented. Existing studies have largely focused on tertiary or referral hospitals, specific professional cadres, or individual facilities, offering limited insight into HH performance across diverse HCW cadres in lower-level district health facilities, where much of the population receives care (1). Moreover, individual-and facility-level factors associated with adequate HH knowledge and good HH practices in these settings remain poorly understood (1, 22). Assessing self-reported HH practices and their determinants is particularly important at the district level because such information can identify gaps in HCW knowledge, motivation, confidence, and enabling conditions that may not be apparent from facility-level compliance measures alone. These findings can inform targeted capacity-building, resource allocation, and locally appropriate IPC interventions in resource-constrained district health facilities.

To address these evidence gaps, this cross-sectional study assessed HH knowledge and practices among HCWs and identified individual-and facility-level factors associated with these outcomes in selected HCFs in the Mukono and Kagadi districts, Uganda. Specifically, the study aimed to: (i) determine the level of HH knowledge among HCWs; (ii) determine the level of HH practices among HCWs; and (iii) identify individual-and facility-level factors associated with HH knowledge and practices among HCWs.

## Methods

### Study Design and Setting

This study used a cross-sectional design to assess HH knowledge and practices among HCWs. The study is reported in accordance with the Strengthening the Reporting of Observational Studies in Epidemiology (STROBE) guideline for cross-sectional studies (S1 Checklist). The study was conducted in two districts: Mukono and Kagadi. Mukono district is located in central Uganda, approximately 21.9km from Kampala City. It is one of the largest districts in the country, with an estimated population of 929,224 (26). It has a population density of approximately 179 people per km², and over 87% of the population lives in rural areas. The district has 22 clinics, 69 HC IIs, 16 HC IIIs, 3 HC IVs, and 3 general hospitals (27). Kagadi District is located in Western Uganda, approximately 255 km from Kampala City. The district is located in the midwestern sub-region, with a total population of 471,111 (26), and the district is predominantly rural. Kagadi district has a total of 31 HCFs, of these, 19 are government, 8 PFPs and 4 PNFPs. The district has 4 clinics, 11 HC IIs, 14 HC IIIs, 1 HC IV and one general hospital (27). The inclusion of predominantly rural areas was intentional because rural communities constitute a substantial proportion of the population served by the healthcare system in the selected districts. Studying HCFs in these settings, therefore, provided an opportunity to assess HH knowledge and practices in contexts where healthcare resources, infrastructure, and access to IPC support may differ from those in more urban settings.

Data were collected from 32 selected HCFs, comprising 16 facilities per district. Of these facilities, 21 were public, 4 were private, for-profit (PFP), and 7 were private, not-for-profit (PNFP). The selected HCFs represented different levels of healthcare delivery and included 22 Health Centre (HC) IIIs, 3 HC IVs, 2 clinics, and 5 general hospitals. The distribution of the selected HCFs broadly mirrored the distribution of these facility types across the study districts, ensuring representation of different levels of healthcare delivery. However, facilities were purposively selected for their relatively high patient volumes and for providing routine medical, surgical, and maternity services to ensure adequate numbers of eligible participants and observations. Thus, selection was not based on random sampling of all HCFs in the study districts. More details have also been published elsewhere (1)

### Study Design and Setting

This cross-sectional study was conducted in selected HCFs in the Mukono and Kagadi districts, Uganda. The selected facilities spanned different levels of healthcare delivery and ownership, including 22 HC IIIs, 3 HC IVs, 2 clinics, and 5 general hospitals. The facilities comprised public, private not-for-profit (PNFP), and private for-profit (PFP) facilities and served predominantly rural and peri-urban populations. They provided a range of preventive, promotive, curative, maternal and child health, emergency, and inpatient services. The two districts were selected to provide variation in geographical and health-system contexts. The distribution of selected facilities broadly reflected the range of facility types available in the study districts. However, facilities were purposively selected based on relatively high patient volumes and the provision of routine medical, surgical, and maternity services; therefore, they were not randomly sampled from all HCFs in the two districts. Further details on the study setting have been published elsewhere elsewhere (1)

### Participants, Eligibility, and Sampling

The study population comprised HCWs employed at the selected HCFs. Eligible participants were frontline HCWs involved in patient care or healthcare service delivery, including medical officers, clinical officers, nurses, midwives, nursing assistants, and other relevant cadres. HCWs were eligible if they had worked at the facility for at least 3 months before data collection and were present during the study period. The minimum 3-month employment period ensured sufficient exposure to routine clinical practices and facility-specific IPC and HH procedures. HCWs who were absent during data collection due to leave, training, or other reasons, did not meet the eligibility criteria, or declined to provide informed consent were excluded. A multistage sampling approach was used. After purposive selection of the HCFs, medical, surgical, and maternity departments were included because of their frequent patient contact and relevance to IPC and HH practices. Staff lists and duty rosters were obtained from each participating facility and used to identify eligible HCWs. The required number of HCWs was allocated proportionally by facility staff size, with consideration of cadres and departments involved in direct patient care. Within each facility and department, eligible HCWs were selected using systematic or simple random sampling, depending on the availability and structure of the staff roster.

When a selected HCW declined participation or was unavailable during data collection, a replacement was selected using the same sampling procedure from the remaining eligible HCWs in the same facility, department, and cadre, where applicable. This was done to maintain the predetermined sample allocation. The numbers of HCWs listed, approached, eligible, excluded, and recruited are presented in a participant flow diagram that illustrates the sampling and recruitment process.

**Figure 1.**
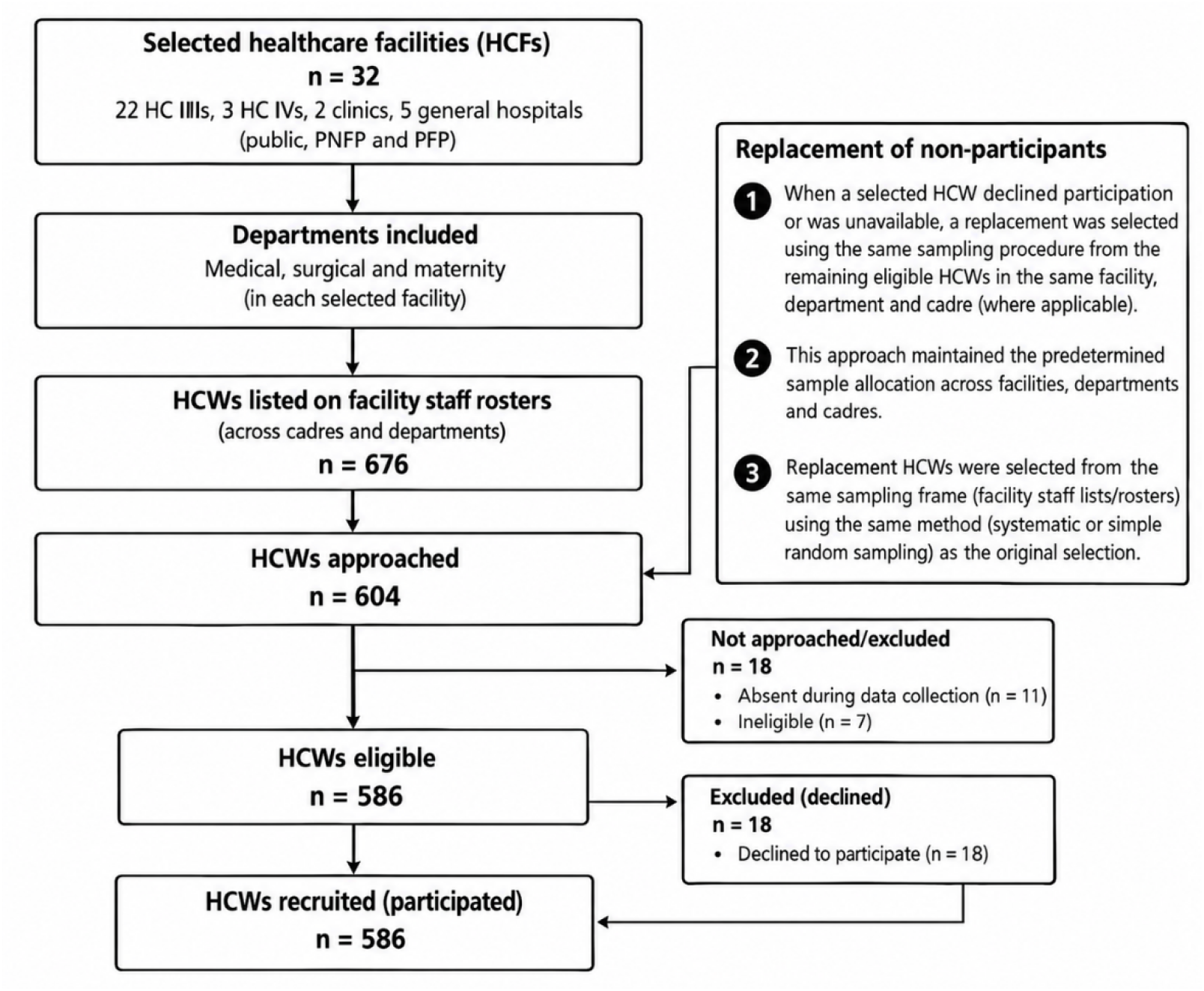
Flow diagram of healthcare worker selection and recruitment

### Sample Size

The initial minimum sample size was calculated using the Kish–Leslie formula for estimating a single population proportion, assuming a 95% confidence level (Z = 1.96), a prevalence (p) of 50% for adequate HH knowledge and practices in the absence of a reliable local estimate, and a 5% margin of error (d), considering the design effect and ICC. The calculation yielded a minimum sample size of 586 HCWs, including a 10% non-response rate. The final sample comprised 586 HCWs. This larger sample was used to achieve the planned allocation across selected HCFs, departments, and HCW cadres and to maximize representation of eligible HCWs within the study facilities. Because HCWs were sampled from 32 HCFs, observations were potentially clustered at the facility level. Accordingly, facility-level clustering was accounted for in the statistical analysis.

### Data collection tools and procedures and Measurement of HH Knowledge and Practice

Data were collected between May and June 2025 from HCWs in selected HCFs in Mukono and Kagadi districts. A structured questionnaire, programmed and preloaded onto the KoboCollect platform, was used to assess HCWs’ HH knowledge and self-reported practices. The questionnaire was developed through a review of relevant literature and existing HH assessment tools and was adapted to the Ugandan healthcare context (28, 29). The questionnaire comprised three sections: socio-demographic and professional characteristics, HH knowledge, and HH practices. The knowledge section assessed HCWs’ understanding of key HH principles, indications for hand hygiene, knowledge on the five moments of HH and IPC measures, while the practice section evaluated self-reported adherence to recommended HH practices during routine patient care and healthcare service delivery. Data were collected through face-to-face interviews conducted by trained research assistants in private locations within the participating HCFs to ensure confidentiality and minimize disruptions to routine clinical activities. The questionnaire was administered in English, the official language of healthcare service delivery in Uganda. Where necessary, questions were clarified in the local language (*Luganda* in Mukono district and *Runyoro* in Kagadi district) to facilitate comprehension and accurate responses. The questionnaire was pretested among HCWs at a health facility with characteristics similar to those of the study sites but located outside the study area. Findings from the pretest informed revisions to improve the clarity, relevance, and consistency of the data collection tool. Questionnaires were administered for approximately 20–30 minutes.

HCWs’ HH knowledge was assessed with a structured questionnaire adapted from the World Health Organization (WHO) HH guidelines and prior IPC studies. The assessment focused on five key knowledge domains: (i) the recommended duration for handwashing with soap and water (>40-60 seconds), (ii) the most effective method for preventing HAIs (proper HH), (iii) situations in which ABHR is not recommended (visibly soiled hands), (iv) the minimum effective alcohol concentration for ABHR (≥75-80%), and (v) complete recall of the WHO Five Moments for HH (before touching a patient, before an aseptic procedure, after body fluid exposure, after touching a patient, and after touching a patient’s surroundings). A composite knowledge score was calculated, and HCWs were classified as having adequate HH knowledge if all five items were answered correctly; otherwise, they were classified as having inadequate HH knowledge. HH practice was assessed by self-reported adherence to recommended HH behaviors during routine patient care. Respondents reported how often they performed HH after patient contact, before aseptic procedures, and used ABHR when soap and water were unavailable. Responses were recorded on a five-point Likert scale from “always” to “never.” Participants also rated their overall compliance with HH guidelines. A composite practice outcome was constructed from four indicators: (i) always performing HH after patient contact, (ii) always performing HH before aseptic procedures, (iii) always using ABHR when soap and water were unavailable, and (iv) self-rating HH compliance as “good” or “excellent.” HCWs who met at least three of the four criteria were classified as having good HH practice, while those meeting fewer than three criteria were classified as having poor HH practice.

### Quality control and assurance

Regular supervision and field monitoring were conducted throughout the data collection period to ensure adherence to study procedures and maintain high-quality data. Additionally, data collection was conducted by trained research assistants with prior experience in health research and electronic data collection. All research assistants held at least a bachelor’s degree in a health-related field, and some held postgraduate qualifications in public health. Before data collection, the research assistants completed a comprehensive three-day training program led by the study investigators. The training covered the study objectives, research ethics, informed consent procedures, questionnaire administration, interviewing techniques, the use of the KoboCollect platform, and data quality assurance procedures. Mock interviews and field simulations were conducted to standardize data collection practices and ensure consistency among data collectors. To minimize data entry errors, the questionnaire was configured in Kobo Collect with built-in validation checks, skip patterns, and mandatory response fields. Throughout the data collection period, field supervisors and study investigators conducted daily reviews of submitted questionnaires to assess completeness, accuracy, and internal consistency. Any discrepancies or missing information identified during these reviews were promptly communicated to the research assistants for clarification and correction. Regular field supervision and monitoring visits were conducted to ensure adherence to study protocols and standardized data collection procedures. Data were uploaded daily to a secure, password-protected server, with access restricted to authorized study personnel. All electronic data were routinely backed up to safeguard against data loss and to maintain data security, confidentiality, and integrity throughout the study.

## Data management and analysis

Completed questionnaires were uploaded daily via the KoboCollect platform and then exported to Microsoft Excel for data cleaning and quality checks. The cleaned dataset was then imported into Stata version 14 for data management and statistical analysis. Categorical variables were coded numerically prior to analysis. Composite indices were developed to assess HCWs’ knowledge and practice regarding HH. For the knowledge domain, responses to objective HH knowledge questions, including knowledge of the WHO Five Moments for HH, recommended handwashing duration, prevention of HAIs, appropriate use of ABHR, and the recommended alcohol concentration for ABHR, were scored. Correct responses received a score of one (1), while incorrect or “don’t know” responses received a score of zero (0). Similarly, a composite HH practice score was generated from responses to four key self-reported HH practice indicators: hand hygiene after patient contact, hand hygiene before aseptic procedures, self-rated HH adherence (good or excellent), and use of ABHR when soap and water were unavailable. For the three frequency items, only responses of “always” were considered appropriate practice and assigned a score of one (1), while “most of the time,” “sometimes,” “rarely,” and “never” were assigned a score of zero (0). For self-rated compliance, “good” or “excellent” was scored as one (1). HCWs who scored at least 3 out of 4 (≥75%) were categorized as having good HH practice, whereas those scoring less than 3 out of 4 (<75%) were categorized as having poor HH practice. HCWs’ self-perceived socioeconomic status (SES) was assessed using a 10-step social ladder scale and categorized into three levels: low (steps 1–3), middle (steps 4–6), and high (steps 7–10) (30, 31). Descriptive statistics summarized participants’ socio-demographic and professional characteristics, HH knowledge, and HH practices. Categorical variables were presented as frequencies and percentages. Cross-tabulations and Pearson’s chi-square tests were used to assess associations between outcome variables and selected explanatory variables. Since the prevalence of adequate HH knowledge and good HH practice exceeded 10%, modified Poisson regression (log link with robust sandwich standard errors) was estimated using generalized estimating equations (GEE) to identify factors associated with the outcomes and to estimate prevalence ratios (PRs). Variables with a p-value <0.20 in bivariate analysis, along with variables considered important based on existing literature, were included in the multivariable models. To account for clustering of HCWs within HCFs, generalized estimating equations (GEE) with an exchangeable working correlation structure were fitted, with clustering specified at the facility level. Crude PRs were estimated from GEE models containing only the variable of interest, and adjusted PRs from models including all variables shown in Tables 4 and 5. Multicollinearity among independent variables was assessed using variance inflation factors (VIFs), and variables with VIFs exceeding 10 were excluded from the final models. Adjusted prevalence ratios (aPRs) and their corresponding 95% confidence intervals (CIs) were reported to quantify the strength of associations. Statistical significance was assessed at the 5% level (p < 0.05). Model fit and parsimony were evaluated using the quasi-likelihood under the independence model criterion (QIC), with lower QIC values indicating better-fitting models.

## Ethical considerations

Ethical approval was obtained from the Makerere University School of Public Health Research and Ethics Committee (Approval No. SPH-2024-632), and the study was registered with the Uganda National Council for Science and Technology (Registration No. HS5449ES). Administrative clearance was obtained from district health authorities and participating healthcare facilities. Written informed consent was obtained from all participants after they were informed about the study objectives, procedures, potential risks, and benefits. Participation was voluntary, and participants could decline or withdraw at any time without consequences. Confidentiality and privacy were maintained through the use of unique identifiers and restricted access to study data.

## Results

### Background characteristics of respondents

A total of 586 HCWs responded to the questionnaire across 32 health facilities in Mukono and Kagadi Districts, representing a 97% response rate (N=604).A small proportion (3%) did not participate because they declined or were unavailable during the data collection period. More than half (53.6%) of the respondents were from Mukono District, and 64.2% practiced in government/public facilities. Slightly under half (49.5%) were attached to health center (HC) IIIs, followed by hospitals (32.6%). Nurses and nursing assistants comprised the largest cadre at 46.2%, followed by midwives at 24.6%. Most respondents were female (69.1%). The mean age of the HCWs was 31.5 years (SD = 8.1; range 19-65), and the mean years of clinical experience was 7.0 years (SD = 6.2; range 1-43). Diploma holders accounted for 36.9% of the sample, while 48.6% held a certificate-level qualification and 14.5% held a bachelor’s degree. Most respondents were either single (47.3%) or married/cohabiting (50.9%), and Catholics were the majority religious group (37.7%). Just over half (54.9%) resided in an urban setting, and 53.9% self-reported a middle socio-economic position. Medical wards accounted for the majority of practice placements (56.7%), followed by maternity (37.5%). The overwhelming majority of HCWs (97.4%) reported ever receiving HH training, with 98.8% reporting their most recent training within the past year (**Table 1**).

**Table 1.**
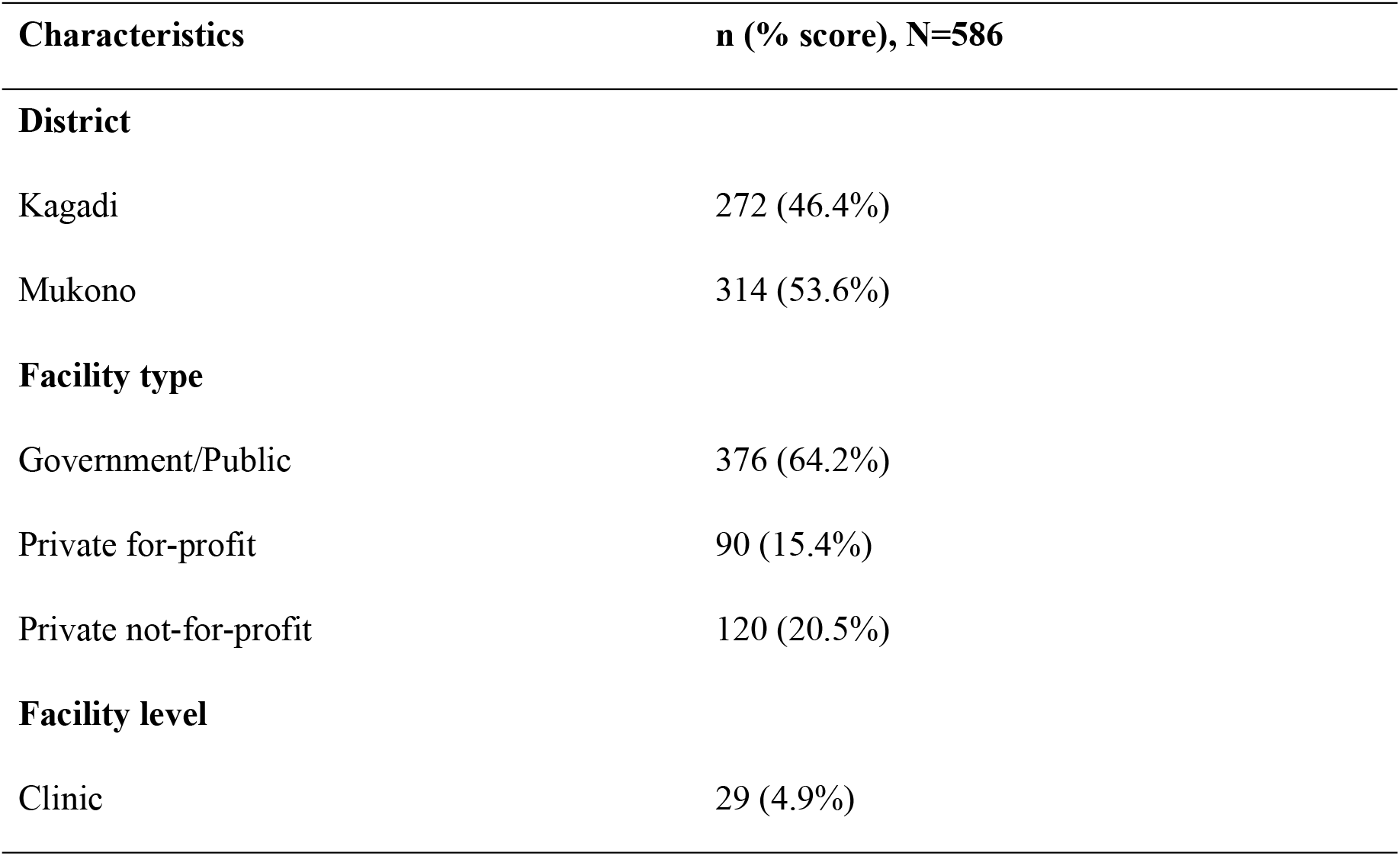
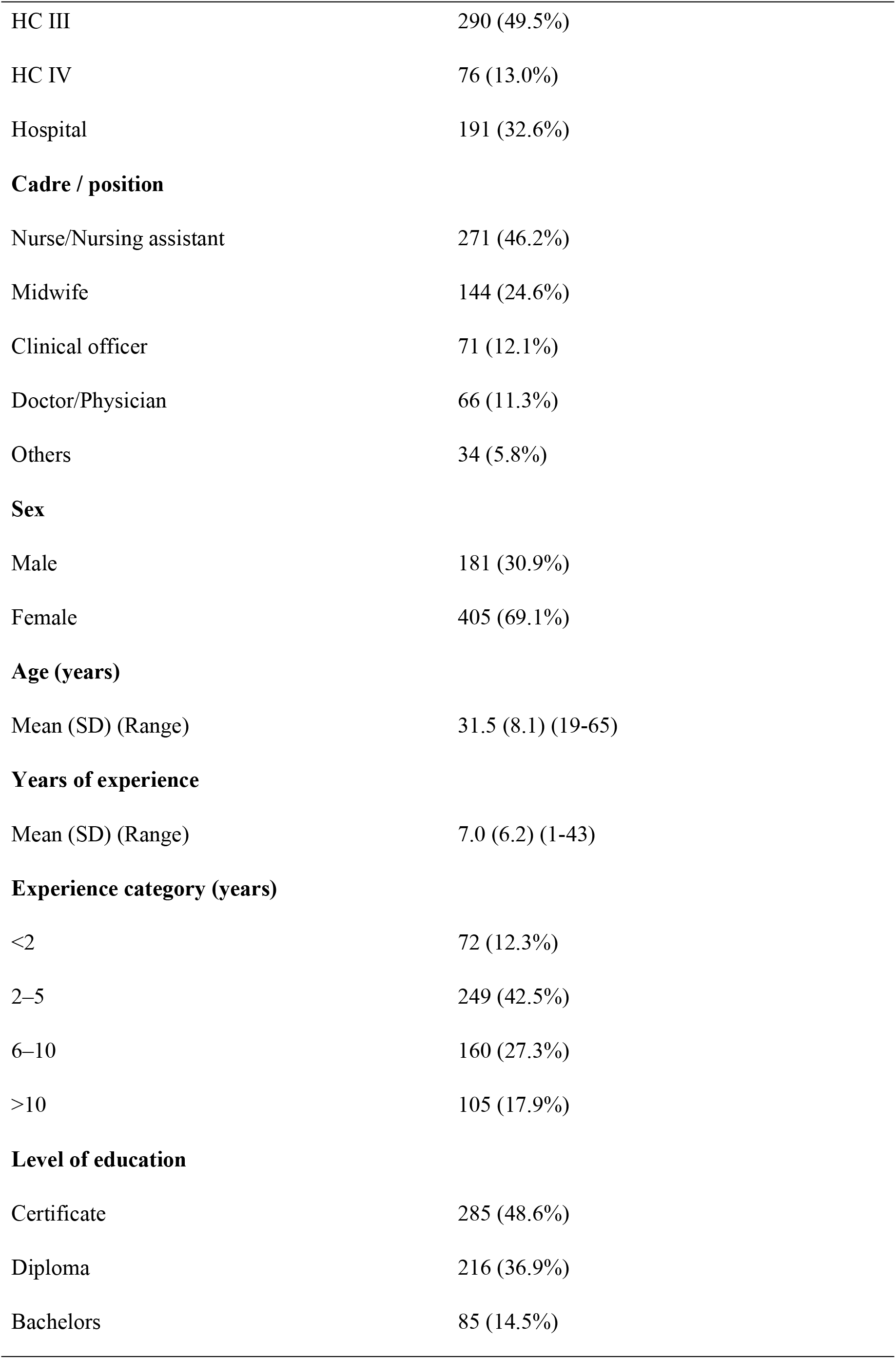
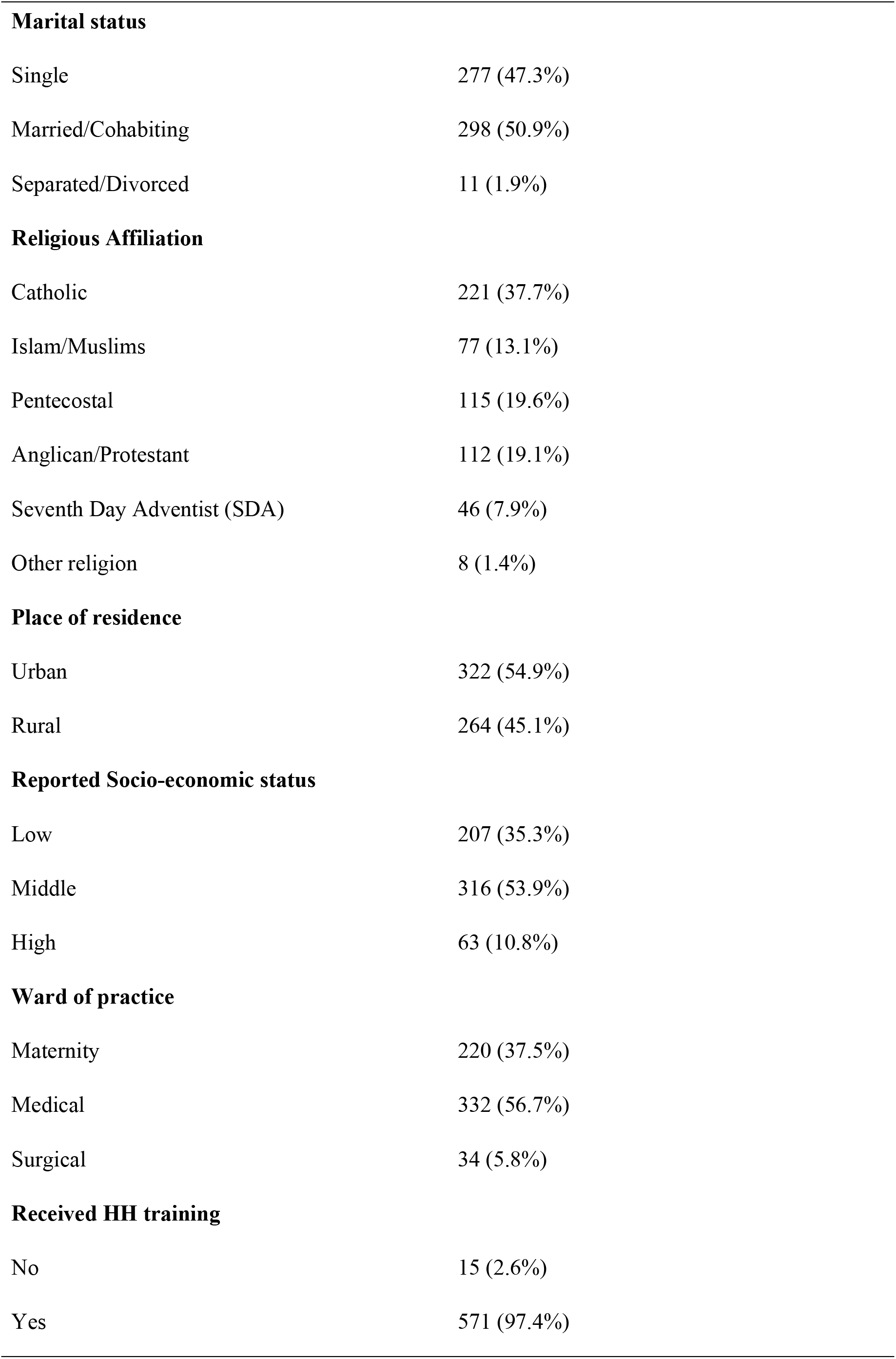
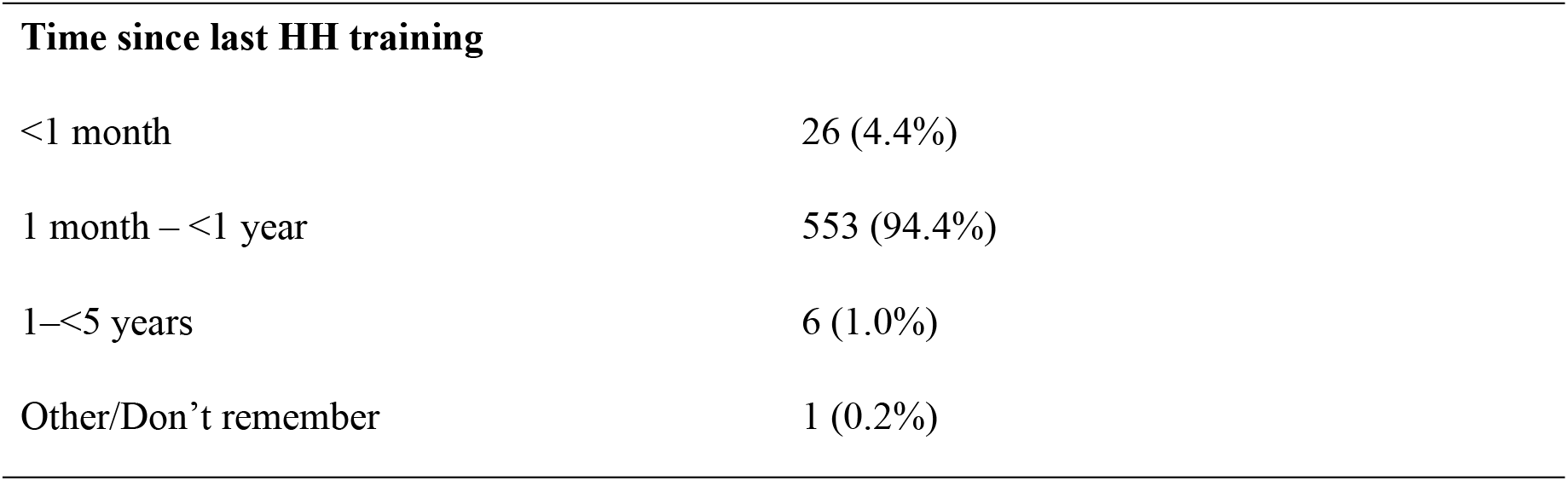
Background characteristics of respondents.

| Characteristics | n (% score), N=586 |
| --- | --- |
| **District** |  |
| Kagadi | 272 (46.4%) |
| Mukono | 314 (53.6%) |
| **Facility type** |  |
| Government/Public | 376 (64.2%) |
| Private for-profit | 90 (15.4%) |
| Private not-for-profit | 120 (20.5%) |
| **Facility level** |  |
| Clinic | 29 (4.9%) |

| **Characteristics** | **n (% score), N=586** |
| --- | --- |
| HC III | 290 (49.5%) |
| HC IV | 76 (13.0%) |
| Hospital | 191 (32.6%) |
| **Cadre / position** |  |
| Nurse/Nursing assistant | 271 (46.2%) |
| Midwife | 144 (24.6%) |
| Clinical officer | 71 (12.1%) |
| Doctor/Physician | 66 (11.3%) |
| Others | 34 (5.8%) |
| **Sex** |  |
| Male | 181 (30.9%) |
| Female | 405 (69.1%) |
| **Age (years)** |  |
| Mean (SD) (Range) | 31.5 (8.1) (19-65) |
| **Years of experience** |  |
| Mean (SD) (Range) | 7.0 (6.2) (1-43) |
| **Experience category (years)** |  |
| <2 | 72 (12.3%) |
| 2–5 | 249 (42.5%) |
| 6–10 | 160 (27.3%) |
| >10 | 105 (17.9%) |
| **Level of education** |  |
| Certificate | 285 (48.6%) |
| Diploma | 216 (36.9%) |
| Bachelors | 85 (14.5%) |
| **Marital status** |  |
| Single | 277 (47.3%) |
| Married/Cohabiting | 298 (50.9%) |
| Separated/Divorced | 11 (1.9%) |
| **Religious Affiliation** |  |
| Catholic | 221 (37.7%) |
| Islam/Muslims | 77 (13.1%) |
| Pentecostal | 115 (19.6%) |
| Anglican/Protestant | 112 (19.1%) |
| Seventh Day Adventist (SDA) | 46 (7.9%) |
| Other religion | 8 (1.4%) |
| **Place of residence** |  |
| Urban | 322 (54.9%) |
| Rural | 264 (45.1%) |
| **Reported Socio-economic status** |  |
| Low | 207 (35.3%) |
| Middle | 316 (53.9%) |
| High | 63 (10.8%) |
| **Ward of practice** |  |
| Maternity | 220 (37.5%) |
| Medical | 332 (56.7%) |
| Surgical | 34 (5.8%) |
| **Received HH training** |  |
| No | 15 (2.6%) |
| Yes | 571 (97.4%) |
| **Time since last HH training** |  |
| <1 month | 26 (4.4%) |
| 1 month – <1 year | 553 (94.4%) |
| 1–<5 years | 6 (1.0%) |
| Other/Don't remember | 1 (0.2%) |

### Distribution of HCWs’ knowledge on various HH aspects

The overall proportion of HCWs with adequate HH knowledge, defined as correctly answering all five knowledge items, was 82.1%, with a higher proportion in Mukono (93.3%) than in Kagadi (69.1%) (p < 0.001). Nearly all respondents (98.8%) correctly identified the recommended hand washing duration as more than 30 seconds. Most HCWs (97.6%) recognized proper HH as the most effective method for preventing HAIs. Additionally, 89.9% correctly stated that ABHR is not recommended when hands are visibly soiled, while 98.8% correctly identified the recommended minimum alcohol content of ABHR as 75-80%. Knowledge of the WHO Five Moments for HH was also high, with 89.9% of respondents correctly listing all five moments (**Table 2**).

**Table 2.** Distribution of HCWs’ knowledge on HH.

| Characteristic | n (% score) |
| --- | --- |
| **Overall adequate Knowledge** | 481 (82.1%) |
| **District** |  |
| Kagadi | 188 (69.1%) |
| Mukono | 293 (93.3%) |
| **Hand-washing duration recommended (seconds)** |  |
| 40-60 | 579 (98.8%) |
| 20-30 | 2 (0.3%) |
| 10-20 | 1 (0.2%) |
| <10 | 2 (0.3%) |
| Not sure | 2 (0.3%) |
| **Most effective method for preventing HAIs** |  |
| Proper hand hygiene | 572 (97.6%) |
| Use of personal protective equipment (PPE) | 11 (1.9%) |
| Regular cleaning of the healthcare environment | 3 (0.5%) |
| **When ABHR is NOT recommended for use** |  |
| Mentioned visibly soiled (correct) | 527 (89.9%) |
| Other response (incorrect) | 59 (10.1%) |
| **Minimum alcohol content in ABHR** |  |
| 75-80 | 579 (98.8%) |
| Lower / other | 7 (1.2%) |
| **WHO 5 Moments for Hand Hygiene** |  |
| All 5 moments listed correctly | 527 (89.9%) |
| Did not list all 5 moments | 59 (10.1%) |

### HCWs’ HH practices

Overall, 43.0% of HCWs demonstrated good HH practice, defined as meeting at least 3 of the 4 optimal HH indicators. Good HH practice was more common among HCWs in Mukono (46.2%) than in Kagadi (39.3%), although the difference was not statistically significant (p = 0.095). Most HCWs reported consistently performing HH after patient contact, with 72.2% indicating they always practiced HH and 23.2% doing so most of the time. Practice of HH before aseptic procedures was lower, although 92.3% reported performing HH either always (45.7%) or most of the time (46.6%). Similarly, 90.8% reported always or most often using ABHR when soap and water were unavailable, while 49.0% reported always doing so. Self-rated compliance with HH guidelines was generally high, with 91.6% rating their compliance as good or excellent. Most HCWs (91.1%) reported that HH was prioritized within their departments, and 48.8% indicated that HH audits or observations were conducted at least monthly (weekly 19.3%; monthly 29.5%). Despite these positive practices, 15.2% of HCWs acknowledged that they did not comply with HH at all critical moments (**Table 3**).

**Table 3.** Description of HCWs’ HH practice.

| **Characteristic** | **n (% score)** |
| --- | --- |
| **Overall, good HH practice** | 252 (43.0%) |
| **District** |  |
| Kagadi | 107 (39.3%) |
| Mukono | 145 (46.2%) |
| **Frequency of HH after patient contact** |  |
| Always | 423 (72.2%) |
| Most of the times | 136 (23.2%) |
| Sometimes | 26 (4.4%) |
| Rarely | 1 (0.2%) |
| **Frequency of HH before aseptic procedure** |  |
| Always | 268 (45.7%) |
| Most of the times | 273 (46.6%) |
| Sometimes | 42 (7.2%) |
| Rarely | 3 (0.5%) |
| **Use of ABHR when soap/water is unavailable** |  |
| Always | 287 (49.0%) |
| Most of the times | 245 (41.8%) |
| Sometimes | 43 (7.3%) |
| Rarely | 6 (1.0%) |
| Never | 5 (0.9%) |
| **Self-rated compliance with HH guidelines** |  |
| Excellent | 142 (24.2%) |
| Good | 395 (67.4%) |
| Average/Fair | 47 (8.0%) |

| Characteristic | n (% score) |
| --- | --- |
| Poor | 2 (0.3%) |
| **HH prioritized in your department** |  |
| Yes | 534 (91.1%) |
| No | 52 (8.9%) |
| **Frequency of HH audits/observations** |  |
| Weekly | 113 (19.3%) |
| Monthly | 173 (29.5%) |
| Quarterly | 128 (21.8%) |
| Rarely | 120 (20.5%) |
| Never | 52 (8.9%) |
| **HCWs comply with HH at all critical moments (self-report)** |  |
| Yes | 497 (84.8%) |
| No | 89 (15.2%) |

**Table 4.**
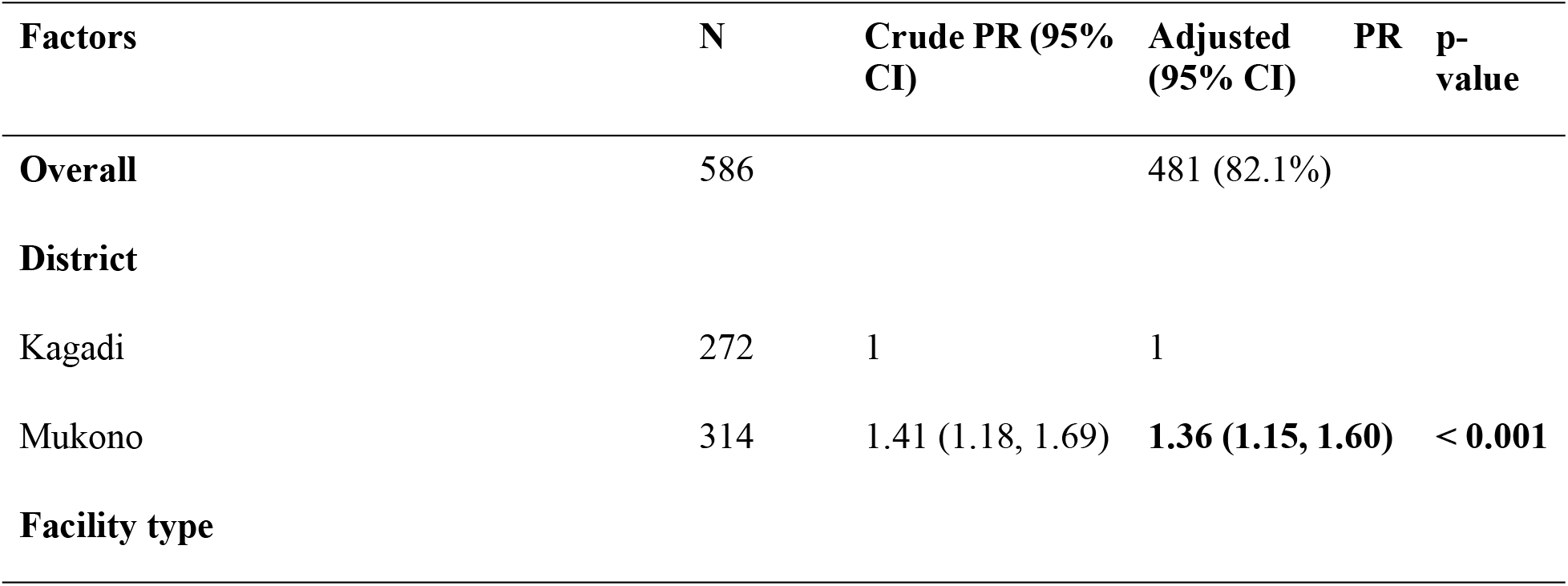
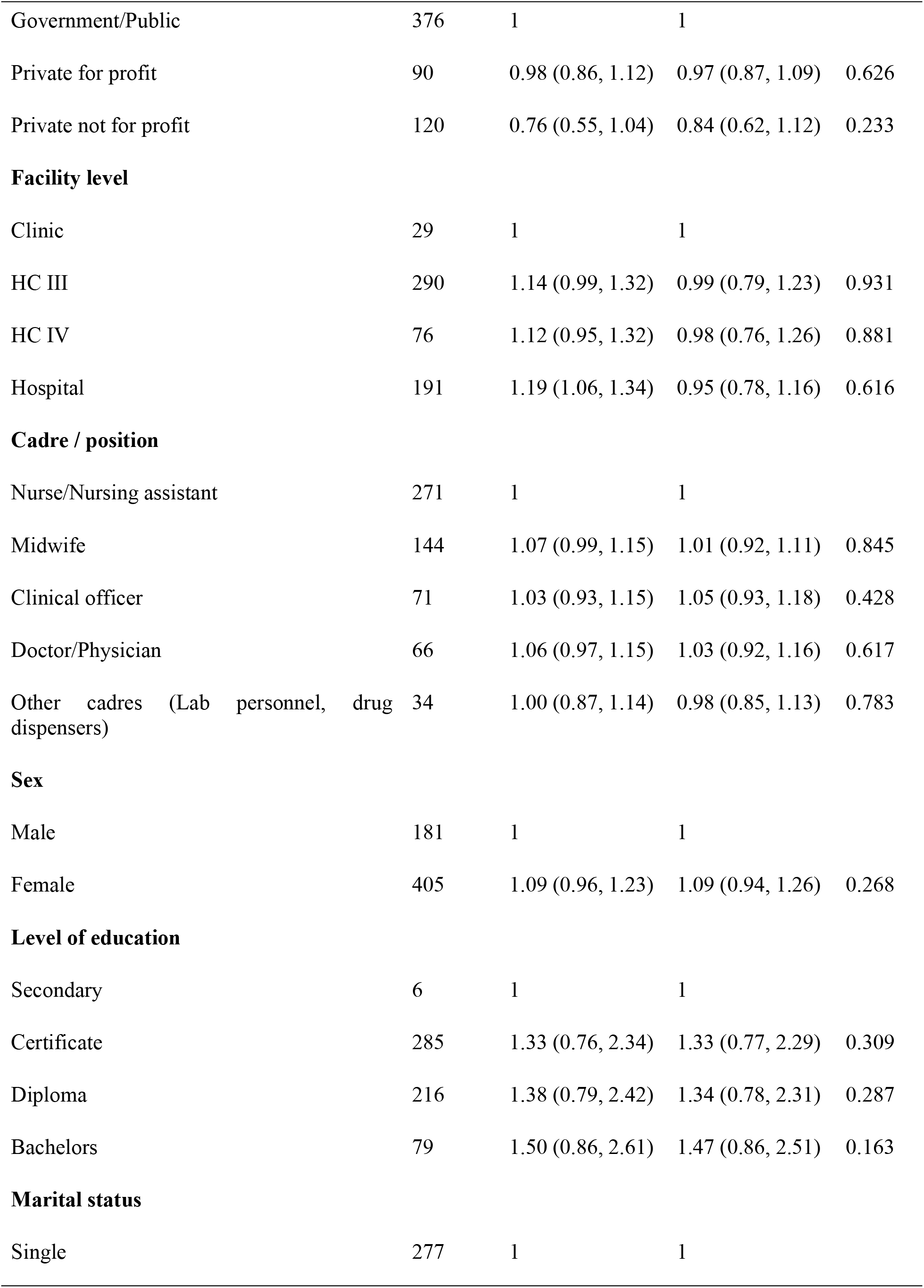
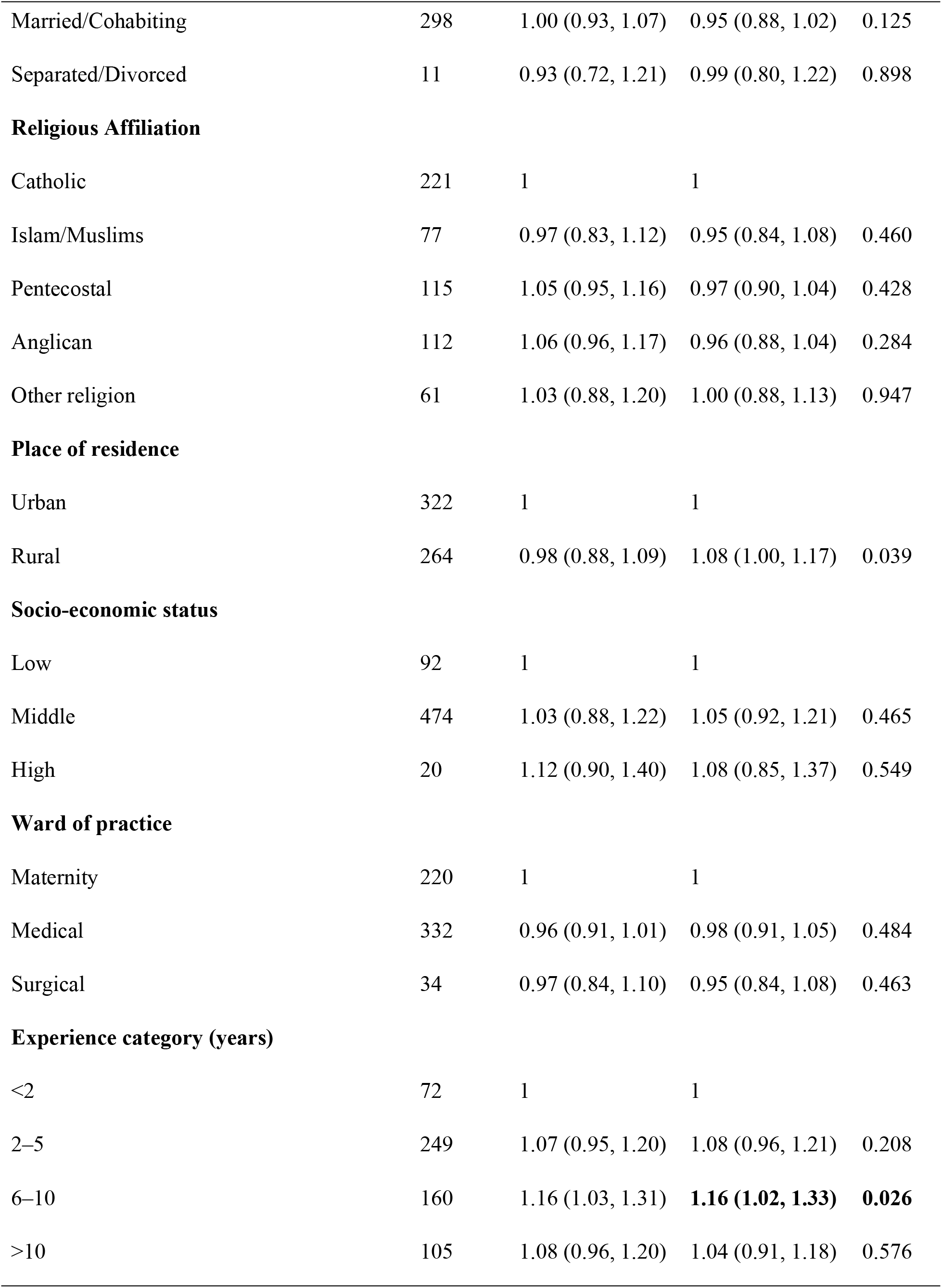
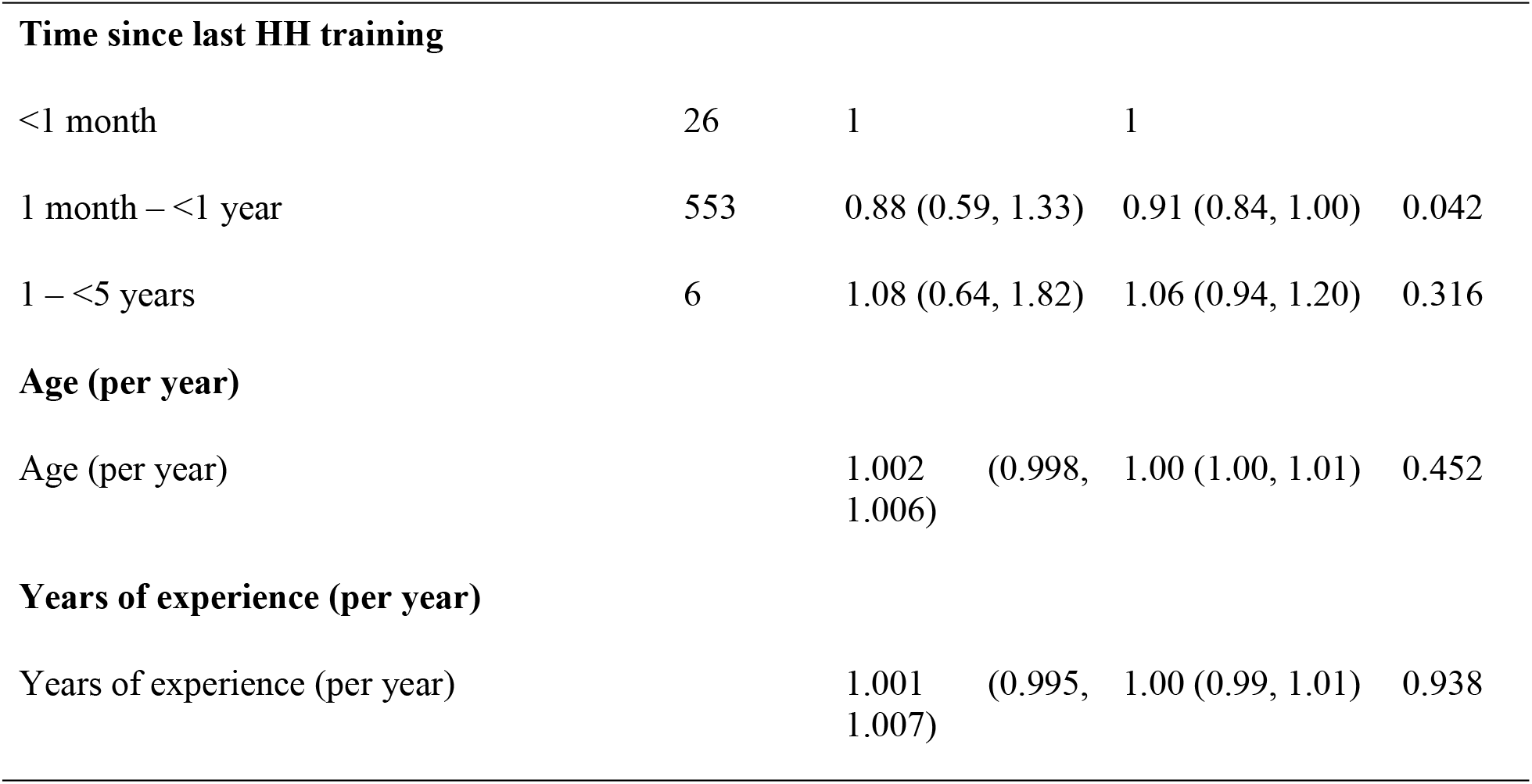
Modified Poisson regression (GEE) of factors associated with adequate HH knowledge among HCWs in Mukono and Kagadi Districts.

**Table 5.**
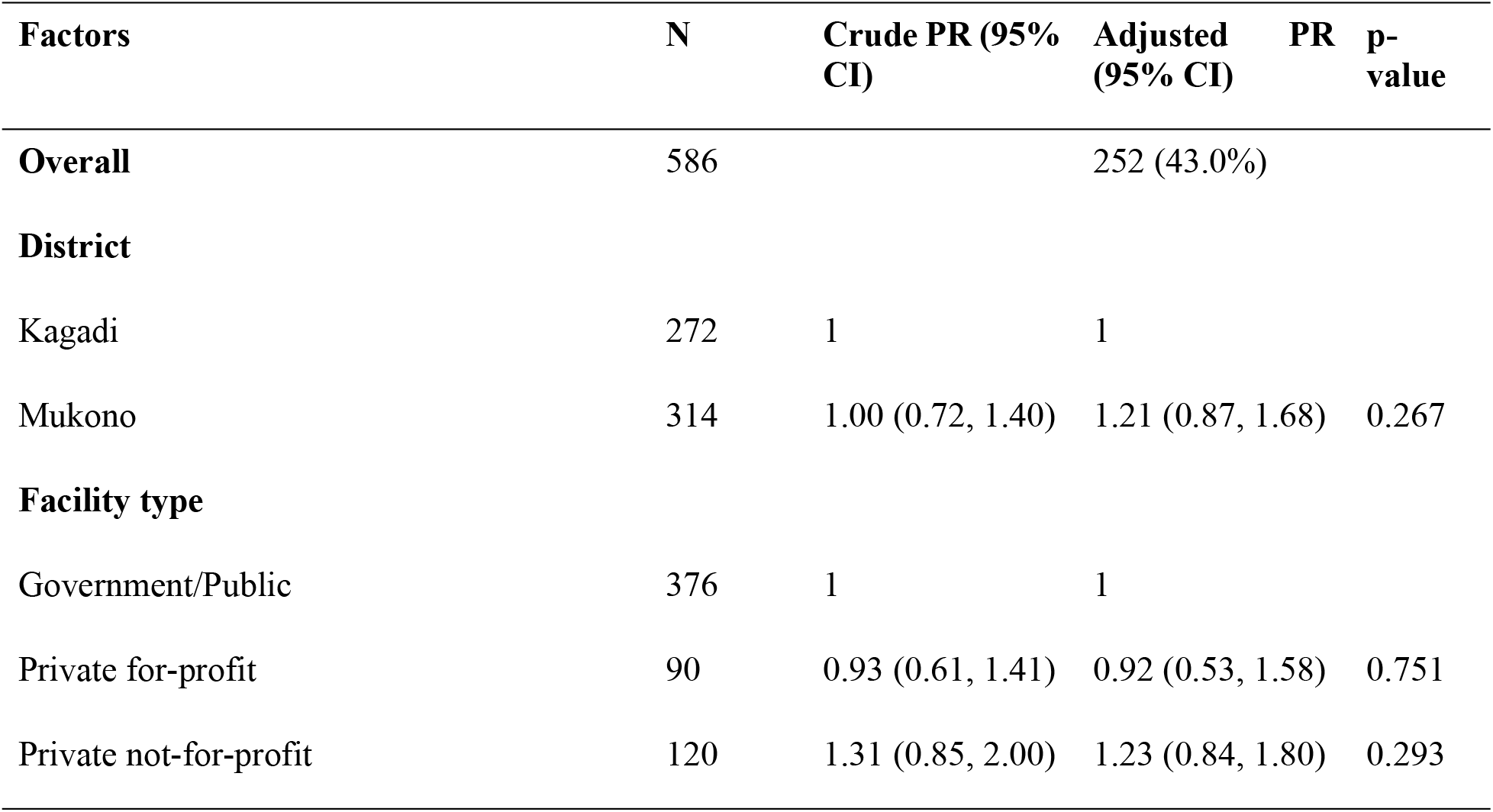
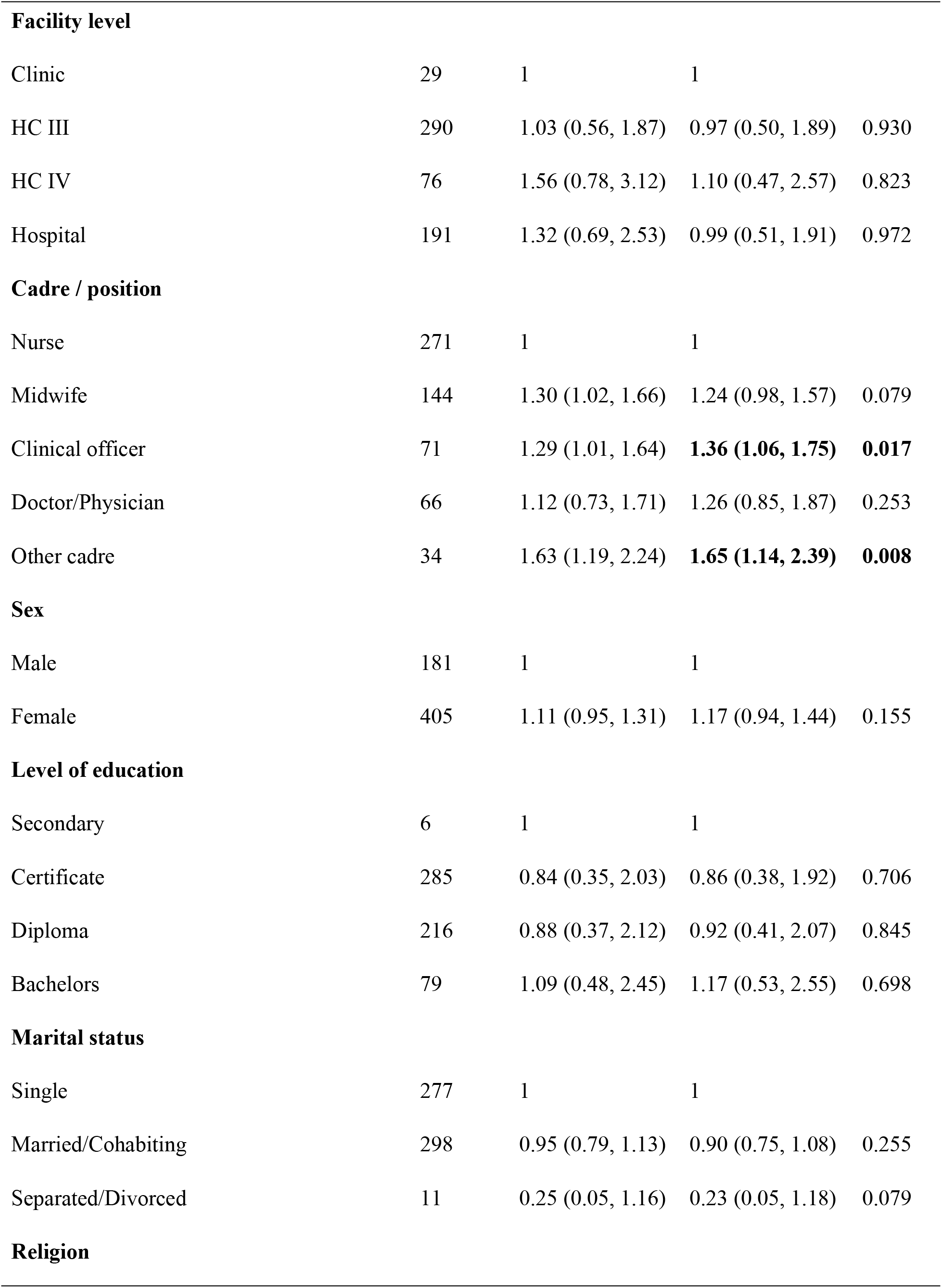
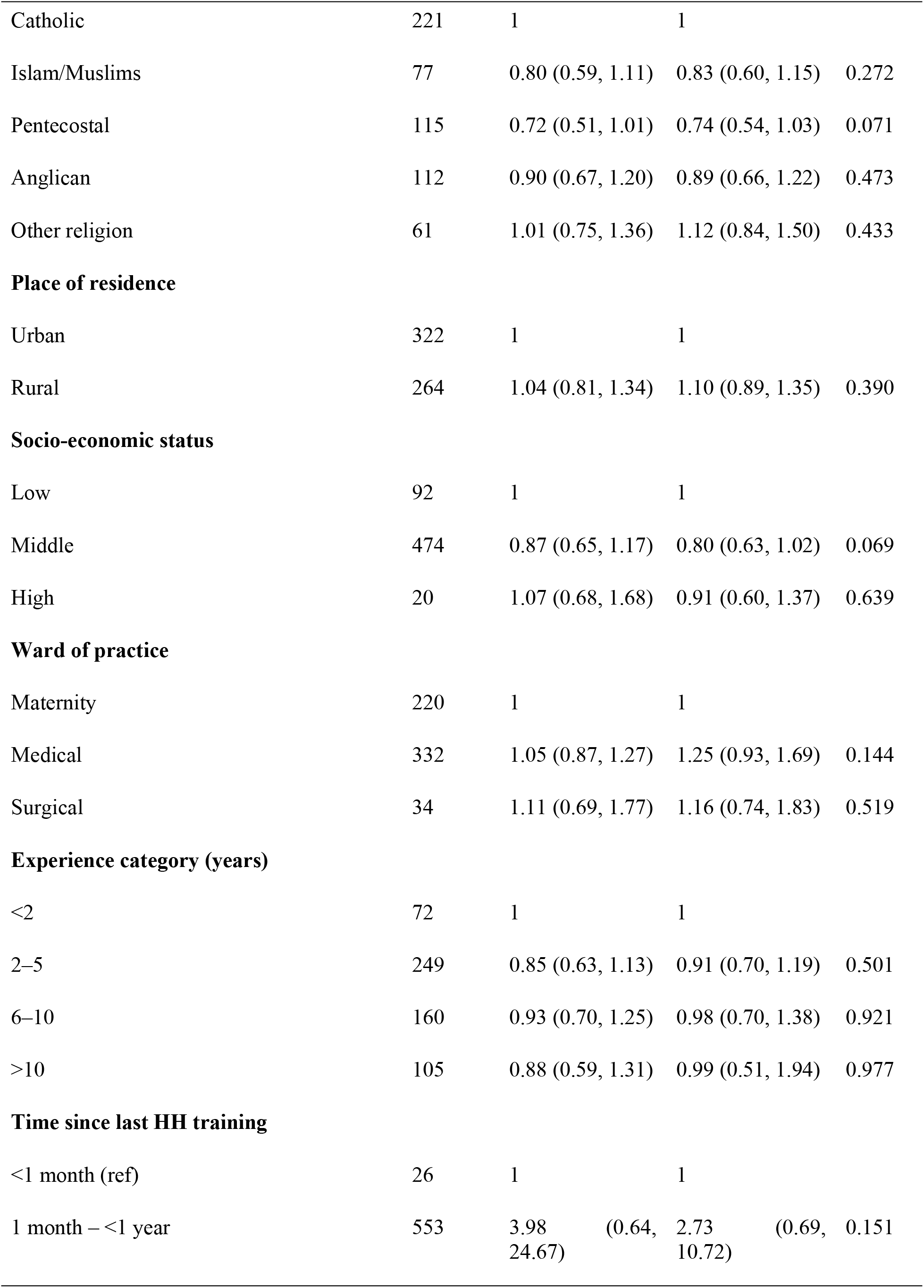
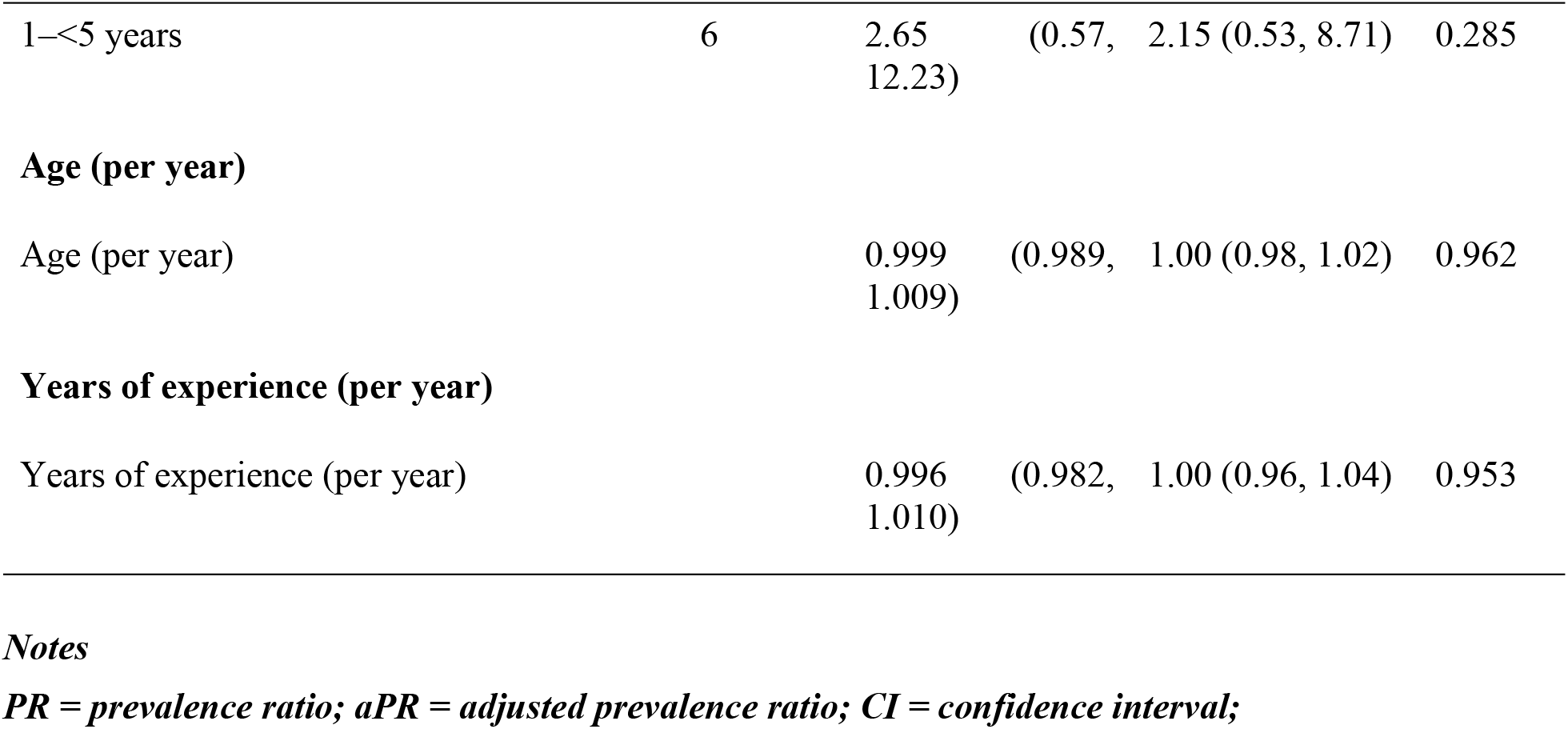
Modified Poisson regression (GEE) of factors associated with good HH practice among HCWs in Mukono and Kagadi Districts.

### Factors associated with the possession of adequate HH knowledge

Bivariate analysis (Pearson’s chi-square test) showed that HCWs from Mukono District had significantly higher HH knowledge than those from Kagadi (p < 0.001). Significant associations were also observed for facility level, work experience, place of residence, and time since the last HH training (all p < 0.05). These variables, together with other variables meeting the pre-specified screening criterion (p < 0.20) and those considered epidemiologically relevant, were entered into the multivariable modified Poisson regression model.

In the multivariable modified Poisson (GEE) regression analysis, several factors were associated with adequate HH knowledge among HCWs, but only a few remained significant after adjustment. HCWs in Mukono District had a 36% higher prevalence of adequate HH knowledge than those in Kagadi District (aPR = 1.36, 95% CI: 1.15–1.60; p < 0.001). Additionally, HCWs with 6-10 years of work experience had a 16% higher prevalence of adequate HH knowledge than those with less than two years of experience (aPR = 1.16, 95% CI: 1.02–1.33; p = 0.026). HCWs in rural areas also had a modestly higher prevalence of adequate HH knowledge than those in urban areas (aPR = 1.08, 95% CI: 1.00–1.17; p = 0.039). In contrast, facility type, facility level, cadre, sex, education level, marital status, religion, socioeconomic status, ward of practice, continuous age and years of experience were not independently associated with adequate HH knowledge after adjustment. HCWs whose last HH training was 1 month to <1 year earlier had a marginally lower prevalence of adequate knowledge than those trained within the past month (aPR = 0.91, 95% CI: 0.84–1.00; p = 0.042), although this comparison rests on a small reference group (n = 26) (**Table 4**).

### Factors associated with good HH practice

Bivariate analysis indicated that cadre/position was significantly associated with good HH practice, with clinical officers, midwives, and other cadres demonstrating higher proportions of good practice than nurses/nursing assistants. Borderline associations were also observed for religion, marital status, socioeconomic status, sex, ward of practice, and time since the last hand hygiene training (all p < 0.20). These variables, together with other epidemiologically important variables, were included in the multivariable modified Poisson regression analysis.

In the multivariable modified Poisson (GEE) regression analysis, cadre was the only factor independently associated with good HH practice. Compared with nurses and nursing assistants, clinical officers had a 36% higher prevalence of good HH practice (aPR = 1.36, 95% CI: 1.06–1.75; p = 0.017), while HCWs in other cadres had a 65% higher prevalence (aPR = 1.65, 95% CI: 1.14–2.39; p = 0.008). Midwives also had a higher prevalence of good HH practice than nurses and nursing assistants, although the association did not reach statistical significance (aPR = 1.24, 95% CI: 0.98–1.57; p = 0.079). [No significant associations were observed between good HH practice and district, facility type, facility level, sex, education level, marital status, religion, place of residence, socioeconomic status, ward of practice, work experience, age, or time since last HH training (**Table 5**).

## Discussion

This study assessed HH knowledge and practices among HCWs in selected HCFs in Mukono and Kagadi districts of Uganda. Overall, the findings show high HH knowledge but substantially lower rates of optimal HH practice. Most HCWs (>80%) had adequate knowledge of HH, while only 43.0% demonstrated good HH practices. This knowledge-practice gap indicates that although HCWs are generally aware of recommended HH principles, translating that knowledge into consistent practice remains a significant challenge. These findings are important as effective HH is widely recognized as the single most effective measure for preventing HAIs and reducing the transmission of antimicrobial-resistant pathogens in HCF settings. The observed knowledge-practice gap highlights a critical implementation challenge for IPC programs in Uganda, particularly at district-level HCFs, where resource constraints may hinder the translation of knowledge into recommended HH practices. Inadequate or inconsistent availability of HH supplies, such as ABHR, soap, and water, may limit HCWs’ ability to consistently apply their knowledge in routine clinical practice, even when they understand the importance of HH (32, 33).

Beyond Uganda, similar gaps in HH compliance threaten patient safety globally, especially in low-and middle-income countries, where HAIs and AMR continue to impose a disproportionate burden on health systems.

The high level of HH knowledge observed in this study (82.1%) may partly reflect HCWs’ widespread exposure to HH training, as nearly all respondents (97.4%) reported receiving HH training (34). These findings suggest that HH training programs implemented by the Ministry of Health, development partners, and HCFs may have successfully disseminated key HH messages. Similar levels of knowledge have been reported in studies conducted in other low-and middle-income countries where regular IPC training and supportive supervision have been institutionalized (34–37). The findings, therefore, suggest that inadequate HH compliance among HCWs in these settings is less likely to be driven by a lack of knowledge and more by challenges in translating knowledge into consistent practice.

Despite high levels of HH knowledge, HCWs demonstrated suboptimal HH practice, highlighting a gap between knowledge and behavior. This knowledge-practice gap has been reported in numerous HH studies worldwide and underscores the complexity of behavior change in healthcare settings(38–41). Knowledge alone is often insufficient to ensure adherence to recommended HH practices (42). Factors such as workload, staff shortages, inadequate access to HH supplies, competing clinical priorities, workplace culture, and weak accountability mechanisms can limit adherence even among knowledgeable HCWs (33, 43). In the present study, HCWs most consistently reported HH after patient contact (72.2% always), whereas consistent HH before aseptic procedures (45.7% always) and consistent ABHR use when soap and water were unavailable (49.0% always) were less common (44). This pattern aligns with evidence from both high-income and LMIC settings, which show that HH adherence tends to be higher during self-protective moments, particularly after patient contact, than during patient-protection moments such as before patient contact or before aseptic procedures (45–47). Such differences may reflect risk perception, with HCWs potentially perceiving the threat of infection after contact with patients as more immediate and personally relevant than the less visible risk of transmitting pathogens to patients before care (48, 49). Similarly, evidence from LMICs further suggests that consistent HH practice, particularly ABHR use, can be constrained by supply availability, point-of-care accessibility, workload pressures, and habitual reliance on soap and water (50, 51). Therefore, the findings suggest that improving HH practice requires interventions that go beyond knowledge enhancement. Such interventions should combine continuous availability and convenient placement of HH supplies with behavioral strategies, supportive supervision, feedback, workplace leadership, and accountability mechanisms to reinforce HH at the point of care. Strengthening these system-and behavior-level components may help translate existing HH knowledge among HCWs into sustained practice.

In addition, WHO-based observational data and multi-country studies have shown that “before aseptic procedure” is among the lowest-performing HH moments globally, despite being one of the most critical for preventing HAIs, particularly surgical site and bloodstream infections (52). Similar findings from studies in sub-Saharan Africa and other LMIC settings highlight that compliance gaps at this moment are often compounded by workload pressures, interruptions in clinical workflow, and inconsistent access to ABHRs at the point of care (33, 44). Collectively, these findings reinforce that HH behavior is not only knowledge-dependent but also strongly influenced by behavioral psychology and health system constraints. Therefore, targeted interventions should go beyond awareness-raising to explicitly reinforce patient-safety-oriented HH behaviors, strengthen the availability of HH resources at the point of care, and integrate behavioral nudges and supervision mechanisms to improve compliance before aseptic procedures.

Nearly 90% of HCWs correctly listed all five WHO “Moments for Hand Hygiene,” indicating a notable level of theoretical knowledge and suggesting successful dissemination of IPC guidelines within the study settings. Similarly high levels of awareness of the WHO Five Moments have been reported in studies from other low-and middle-income countries, as well as in selected high-income settings, where structured IPC training programs and repeated exposure to WHO guidelines have contributed to improved cognitive understanding of HH indications (53, 54). However, despite this strong knowledge base, the observed gap between knowledge of the WHO Moments and actual HH practice underscores a persistent, well-documented challenge in IPC (55–58). Evidence from multiple studies across sub-Saharan Africa and other resource-limited settings consistently shows that high HH knowledge does not automatically translate into high HH compliance (59, 60). This disconnect has been attributed to a complex interplay of factors beyond awareness, including limited availability of alcohol-based hand rubs at the point of care, competing clinical demands, high patient-to-healthcare worker ratios, and workflow interruptions that make adherence to all HH moments difficult in real-time practice (61, 62). In addition, organizational factors such as weak supervisory systems, inconsistent performance feedback, and limited reinforcement of IPC standards at the facility level have been shown to reduce sustained compliance even among well-trained HCWs (63). These findings suggest that improving HH compliance requires more than knowledge alone. Instead, effective interventions should integrate continuous on-site mentorship, real-time performance feedback, improved availability of HH resources at the point of care, and stronger institutional leadership to reinforce a culture of safety. This is especially important in district-level health facilities, where systemic constraints are more pronounced, and most patient contacts occur.

An important finding of this study was the significant variation in HH knowledge across districts, with HCWs in Mukono 36% more likely to demonstrate adequate HH knowledge than those in Kagadi. This disparity may reflect differences in access to health system resources, training opportunities, and implementation support. However, because HCWs residing in rural areas had a slightly higher prevalence of adequate knowledge after adjustment, the district difference is unlikely to be explained by urban–rural residence alone, and differences in facility mix, partner support, or data-collection teams between districts cannot be excluded. Mukono’s proximity to Kampala and other urban centers may provide greater access to pre-and in-service training institutions, continuing professional development (CPD) programs, mentorship, supportive supervision, and exposure to national and international IPC initiatives (64, 65).

Urban and peri-urban health facilities are also more likely to collaborate with implementing partners, academic institutions, and research organizations, thereby increasing opportunities for capacity building and the dissemination of evidence-based IPC practices (66, 67). In contrast, HCWs in more rural and underserved districts often face constraints such as limited human resources, inadequate funding, fewer supervision visits, and restricted access to refresher training, all of which may adversely affect knowledge acquisition and retention (68–70). These findings are consistent with previous studies from low-and middle-income countries showing that geographical disparities in access to training and health system support contribute to inequalities in HCWs’ IPC knowledge and practice (71–73). The observed district-level differences, therefore, highlight the need for context-specific strategies that promote equitable access to high-quality IPC education, regular mentorship, supportive supervision, and continuous professional development, particularly in rural and resource-constrained settings, to strengthen HH knowledge and ultimately improve compliance with recommended infection prevention practices.

The association between years of experience and HH knowledge is consistent with a possible role of professional exposure in knowledge acquisition. HCWs with 6-10 years of experience demonstrated significantly higher HH knowledge than those with less than two years of experience. This may reflect repeated participation in IPC training, mentorship, and clinical exposure, as well as routine engagement in quality improvement activities over time. This has been reported elsewhere (74–76). Newer HCWs may still be developing a practical understanding of IPC guidelines and may require additional orientation and mentorship during the early years of service. Strengthening induction programs and targeted mentorship for newly recruited HCWs could therefore help improve HH knowledge and facilitate early adoption of recommended practices (77–79). Interestingly, after adjustment, educational level, facility level, facility ownership, sex, and professional cadre were not independently associated with HH knowledge. This may indicate that core HH messages have reached HCWs across categories, although the limited variability of the knowledge measure and small subgroup sizes (e.g., n = 6 with secondary education) reduce the power to detect differences. The absence of educational disparities may indicate that IPC training programs are reaching HCWs across multiple professional levels and facility types, thereby promoting a common understanding of HH principles.

In contrast to HH knowledge, the professional cadre emerged as the principal determinant of good HH practice. Clinical officers and HCWs in other cadres were significantly more likely to report good HH practice than nurses and nursing assistants. Midwives also tended to practice better HH, though the association did not reach conventional levels of statistical significance. Similar differences in HH compliance across professional groups have been reported in previous studies, with physicians, clinical practitioners, and staff involved in invasive procedures often demonstrating higher adherence to HH recommendations than nursing personnel in some settings (76). Several explanations may account for these findings. Clinical officers and midwives frequently perform procedures requiring strict aseptic techniques, which may heighten awareness of healthcare-associated infection risks and promote stronger adherence to HH protocols (80). Furthermore, procedural training, mentorship, and routine supervision associated with these roles may reinforce compliance with IPC practices.

Nurses and nursing assistants, who made up nearly half of the study population, often face heavier patient workloads and more frequent patient contact, factors consistently identified as barriers to optimal HH compliance in both high-and low-resource settings (20, 32, 81). Heavy workloads, time constraints, staff shortages, and competing clinical priorities can limit opportunities to perform HH at every recommended moment (33, 82). Since nurses provide most direct patient care and have the most frequent patient interactions, targeted interventions such as workload-sensitive HH programs, regular refresher training, supportive supervision, and improved access to HH supplies could substantially improve overall facility-level compliance. These findings underscore the importance of designing cadre-specific IPC interventions that address the unique operational challenges faced by nursing staff.

The lack of association between recent HH training and HH practice should be interpreted cautiously. Because nearly all respondents had been trained within the past year and the reference group was small (n = 26), the study had limited ability to detect an effect of training. Although nearly all respondents had received training, training alone did not independently predict better HH behavior. This finding aligns with evidence that educational interventions, while necessary, are often insufficient when implemented in isolation (83, 84). Sustainable improvements in HH compliance typically require multimodal interventions that combine training with continuous supervision, performance feedback, point-of-care reminders, leadership engagement, regular audits, and reliable availability of HH supplies (85, 86). The WHO Multimodal Hand Hygiene Improvement Strategy emphasizes this integrated approach, recognizing that behavior change requires supportive systems alongside knowledge enhancement (87).

### Strengths and limitations of the study

One major strength of this study is that it included a relatively large sample of HCWs from multiple facilities across two districts in Uganda, thereby enhancing the precision of the estimates within the study settings. Second, participants were recruited from facilities at different levels and ownership categories, allowing assessment of HH knowledge and practice across diverse HCF contexts. Finally, the study assessed both HH knowledge and practice, enabling examination of the important gap between awareness and implementation of recommended HH behaviors. However, with all its strengths, the study still has certain limitations. HH practice was measured by self-report rather than direct observation, which made the findings susceptible to social desirability and recall bias. Self-reported compliance typically overestimates observed compliance substantially, so the true level of good practice is likely lower than reported. Facilities were purposively selected partly for high patient volume, which may have favoured larger or better-resourced facilities and limits generalisability to lower-volume facilities, including HC IIs, which were not sampled. The knowledge index comprised only five items with high correct-response rates, producing a ceiling effect.

## Conclusion

This cross-sectional study assessed HH knowledge and practices among HCWs and examined individual and facility characteristics associated with these outcomes in selected HCFs in the Mukono and Kagadi districts of Uganda. This study found that HCWs had high HH knowledge but suboptimal HH practice, suggesting a gap between knowledge and self-reported practice. HH knowledge was significantly associated with district of practice and years of professional experience, whereas good HH practice was primarily associated with professional cadre, with clinical officers and other cadres demonstrating better compliance than nurses and nursing assistants. HCFs should ensure the continuous availability of ABHRs and handwashing facilities at the point of care, implement regular supportive supervision with structured feedback on HH performance, and strengthen real-time reminders and visual cues in clinical areas to reinforce compliance during critical HH moments. These recommendations, consistent with the WHO multimodal HH improvement strategy, should be complemented by future studies that combine direct observation of HH compliance with facility-level assessments, such as the WHO Hand Hygiene Self-Assessment Framework, to identify the system-level drivers of practice.

## Data Availability

All the data and materials supporting this manuscript are within the paper and the supplementary file. More information can be obtained from the Research and Ethics Committee of the Makerere University School of Public Health. They can be contacted at.

## Acknowledgements

The authors sincerely thank all HCWs who participated in this study for their time and valuable contributions. We are grateful to the District Health Offices of Mukono and Kagadi for their support and collaboration. We also acknowledge the management and staff of the participating health facilities for their cooperation during data collection. Special appreciation goes to the research assistants, whose dedication and commitment were instrumental to the study’s successful implementation. Finally, we thank all individuals and institutions for their support in completing this work.

